# Impact of COVID-19 on healthcare access for Australian adolescents and young adults

**DOI:** 10.1101/2021.12.01.21267121

**Authors:** Md Irteja Islam, Joseph Freeman, Verity Chadwick, Alexandra Martiniuk

## Abstract

**Background:** Access to healthcare for young people is essential to build the foundation for a healthy life. We investigated the factors associated with healthcare access by Australian young adults during and before the COVID-19 pandemic.

**Methods:** We included 1110 youths using two recent data collection waves from the Longitudinal Study of Australian Children (LSAC). Data were collected during COVID-19 in 2020 for Wave 9C1 and before COVID-19 in 2018 for Wave 8. The primary outcome for this study was healthcare access. Both bivariate and multivariate logistic regression models were employed to identify the factors associated with reluctance to access healthcare services during COVID-19 and pre-COVID-19 times.

**Results:** Among respondents, 39.6% avoided seeking health services during the first year of the COVID-19 pandemic when they needed them, which was similar to pre-COVID-19 times (41.4%). The factors most strongly impacting upon reluctance and/or barriers to healthcare access during COVID-19 were any illness or disability, and high psychological distress. In comparison, prior to the pandemic the factors which were significantly impeding healthcare access were country of birth, state of residence, presence of any pre-existing condition and psychological distress. The most common reason reported (55.9%) for avoided seeking care was that they thought the problem would go away.

**Conclusions:** A significant proportion of youths did not seek care when they felt they needed to seek care, both during and before the COVID-19 pandemic.

**What is known about the subject?:**

- Some adolescents and young adults do not access healthcare when they need it.
- Healthcare access and barriers to access is best understood through a multi-system lens including policy, organisational, and individual-level factors. For instance, policy barriers (such as cost), organisational barriers (such as transportation, or difficulty accessing a timely appointment) and individual barriers (such as experiences, knowledge or beliefs).
- Barriers to care may differ for sub-groups e.g. rural
- During the COVID-19 pandemic, public health restrictions including the stricter “lockdowns” have reduced healthcare access. The burden of cases upon the healthcare system has further reduced healthcare access.

**What this study adds?:**

- A significant proportion of youth did not seek healthcare when they felt they needed to seek care, both before (41.4%) and during the first year of the COVID-19 pandemic (39.6%)
- Youth with a disability or chronic condition, asthma and/or psychological distress were more likely to avoid accessing healthcare during COVID-19 times.
- The most common reason for not seeking healthcare when it was felt to be needed was because the youth thought the problem would go away (pre-COVID-19 35.7% of the sample versus during the first year of COVID-19 55.9%)
- During the coronavirus restriction period (“lockdown”) the most common reason for not seeking healthcare when it was felt to be needed was because the youth did not want to visit a doctor during lockdown (21.8%) with the next most common reason being because telehealth was the only appointment option available at the time (8.4%)

## INTRODUCTION

Access to healthcare is key to maintaining health and optimising disease management. Early in the COVID-19 pandemic, adolescents and young adults were identified as vulnerable to the direct and indirect effects of COVID-19 due to reduced access to healthcare.^1^ Health systems have been burdened by waves of COVID-19, resulting in decreased healthcare resources to manage COVID-19 and non-COVID-19 related illnesses.^2^ Access to healthcare has also been impeded by mandated isolation, travel restrictions, lost or reduced income and support, and the perceived risk of COVID-19 to self and vulnerable persons.^3^ In the last two years, large proportions of outpatient and primary healthcare has moved to telehealth.^4^

Access to healthcare has been reduced in both high-income and low- and middle-income countries, resulting in increased mortality and morbidity.^5, 6^ Access to paediatric healthcare has reduced dramatically in countries including China,^7^ USA,^8^ and Germany.^9^ The pandemic has had positive and negative effects on young people’s access to healthcare. For example, non-pharmacological measures implemented in countries worldwide have reduced paediatric respiratory viruses by up to 80% in some locations, though with societal re-opening, transmission of viruses such as respiratory syncytial virus are liable to surge.^10^

The Australian government has stated that it is approaching the “final post-vaccination phase”,^11^ with high vaccination rates, relatively few deaths, and modest economic impact from COVID-19 in comparison to other countries.^12^ Time will tell whether Australia is truly in its ‘final post-vaccination phase’ given the risk of new variants and the emergence of the Omicron variant just this week. If the worst is behind us, recovery will require providing care that was deferred when healthcare access was limited by the pandemic. This includes system-deferred care such as elective surgery, and patient-deferred care in cases of personal concern.^13^ Unfortunately the care deferred during the pandemic will likely mean increased morbidity and mortality from conditions which either have not been treated adequately or diagnosed at all.

Studies have examined health service use during the pandemic in Australia. Victoria, Australia showed reduced hospital presentations among the young and elderly during the pandemic, compared to before. However, healthcare access for urgent conditions at the population-level remained constant.^14^ A study of paediatric health service use in NSW, Australia found significantly lower attendance in the 2020 lockdown for chronic conditions, acute infections, and injury.^15^ After the lockdown, hospital presentations returned to pre-COVID-19 levels, except for mental health presentations which remained 30-55% higher than predicted.^15^

The definition of ‘access to healthcare’ can be divided into ‘having access’ and ‘gaining access’.^16^ A population has access when healthcare services are available in adequate supply. A population gains access when certain barriers are overcome. These can be ‘financial, organisational, social or cultural.’^16, 17^

In Australia, the COVID-19 pandemic has affected young people in multiple ways. Australians aged 20-29 have the highest total proportion of positive cases.^18^ Although COVID-19 is less severe in young people on average,^1^ concerns remain for chance of severe illness, transmission concern, and foregone non-COVID-19 care.^19^ Emergency Department (ED) presentations for young people aged 15-24 dropped 3.9% from 2018-19 to 2019-20, the latter period spanning 1 July 2019 to 30 June 2020.^20^ This decrease during the pandemic contrasts with an average annual increase of 1.7% from 2016-17 to 2018-19.^20^

Young people in aggregate require less healthcare for acute and chronic illnesses than older age groups.^21^ However, health in adolescence and young adulthood sets a lifelong trajectory.^22^ For children with acute problems, access to safe and effective healthcare reduces risk of complications and interruption to physical and mental development. For children with life-long chronic health conditions, optimising management early, which requires access to healthcare, ensures the best outcomes.^23^

Young people’s mental health has been adversely affected during COVID-19, and this is compounded by lack of access to mental healthcare.^20^ In Australia, three quarters of young people’s mental health was worsened by the COVID-19 pandemic.^24^ Decreased access to healthcare is of concern for young people during the pandemic, especially considering their escalating mental health issues.

Prioritising access to safe and desirable healthcare for young people is important, as without such care they can fall behind in physical, psychosocial, and educational development. This study investigates the impact of COVID-19 on healthcare access for young people using data from the Longitudinal Study of Australian Children.

## METHODS

### Data source

We utilized data from Growing Up in Australia: The Longitudinal Study of Australian Children (LSAC) survey. The LSAC is a population-based national cohort study that has collected detailed information biennially since 2004 from two cohorts who in the first wave were a younger B-cohort (aged 0-1) and older K-cohort (aged 4-5) using a multi-stage sampling approach. Data were collected from parents and/or children (when aged 10 years or older) using personal face-to-face interviews by trained interviewers and/or self-reported computer-assisted questionnaires. The details of the LSAC study design, sampling technique and data collection procedures are described elsewhere.^25^

For this current study, we analysed 1110 of the older cohort (now aged 20-21 years). We analysed two recent LSAC waves - Wave 9C1 and Wave 8. Data were collected in between October to December in 2020 for Wave 9C1 with COVID-19 related information (termed ‘COVID-19 times’) and in 2018 before the COVID-19 pandemic for Wave 8 (termed ‘pre-COVID-19 times’).^25^ The analytical sample was restricted to those who completed data on the outcome variable (service access) and exposure variables (e.g., diagnosed with asthma, medical conditions, psychological distress) in both waves. During analysis, ‘Missing values’ and ‘Do not know’ response categories were omitted.

### Outcome variable

The main outcome variable of the study is service access. The following item was directly asked to K-cohort of both waves (Wave 9C1 and Wave 8) to assess healthcare access: *“In the last 12 months, has there been any time when you thought you should get medical care, but you didn’t?”*.

### Exposure variables

We selected independent variables based on previous studies that identified determinants of health service utilization.^26, 27^ Demographic characteristics included sex (male and female), country of birth (overseas and Australia), states (New South Wales, Victoria, Queensland and Others), remoteness (major cities and regional/remote), schooling (technical/others and university/tertiary), employment (unemployed and employed), living with parents (no and yes), family cohesion (poor and strong), socioeconomic status using Index of Relative Socioeconomic Advantage and Disadvantage (IRSAD) quintiles (Q1-most disadvantaged to Q5-most advantaged). The current study also considered health-related factors including: any illness or disability that needed continuous assistance or supervision and lasted for six months or more; self-reported pre-existing medical conditions (e.g., diabetes, anxiety, autism, heart disease, epilepsy, etc); diagnosed with asthma; and psychological distress using Kessler Psychological Distress Scale (K10) score (categorized as low/moderate and high/very high). Further, we included variables related to the COVID-19 pandemic including the coronavirus restriction period (CRP) (also known as “lockdown”) between March-May 2020 that were only available in LSAC Wave 9C1.^25^ COVID-19 related variables included whether the respondent was tested for COVID-19 or not, whether the participants were involved in physical activities during the CRP, employment status during the CRP, whether the youth received any COVID-19 financial support during the CRP, and life difficulty during the COVID-19 restriction period.

### Statistical analysis

Descriptive statistics in terms of frequency (n) and percentages (%) described the characteristics of the sample, distribution of access to services by LSAC Wave 9C1 (COVID-19 times) and Wave 8 (Pre-COVID-19 times), and the reasons given for not accessing healthcare. Chi-square analyses examined the bivariate associations between independent variables and outcome variable (service use). Finally, multivariate logistic regression models were employed to identify the predictors of ‘no service use’ during COVID-19 times (Model I) and pre-COVID-19 times (Model II), respectively. Regression results were presented in the form of adjusted odds ratios (OR) with 95% confidence intervals (CI). All data were weighted to account for LSAC’s multi-clustered study design and analyses were implemented using the ‘svyset’ package of Stata 14.1 version.

## RESULTS

The characteristics of the samples are detailed in Table 1. A total of 1110 youths were selected for the study, using two LSAC waves – W9C1 and W8. The study population included 651 (58.6%) females with mean age 20.63 years. The majority of respondents were born in Australia, nearly 55% of youths were from New South Wales and Victoria combined and the remainder from the other states in Australia, and more than three-fourths (76.5%) were from major cities. Sixty-four percent were enrolled in university or tertiary level education and 77.7% were employed. Overall, 71.5% of young people were living with their parents, almost 85% reported strong family cohesion, and nearly 70% of youths were from disadvantaged socioeconomic groups (Quartiles 1-3 combined). Further, 23.7% of youths had an illness, medical condition or disability (which required continuous assistance), 43% had self-reported pre-existing conditions, nearly one-quarter had asthma and more than 40% had high to very high psychological stress.

**Table 1.**
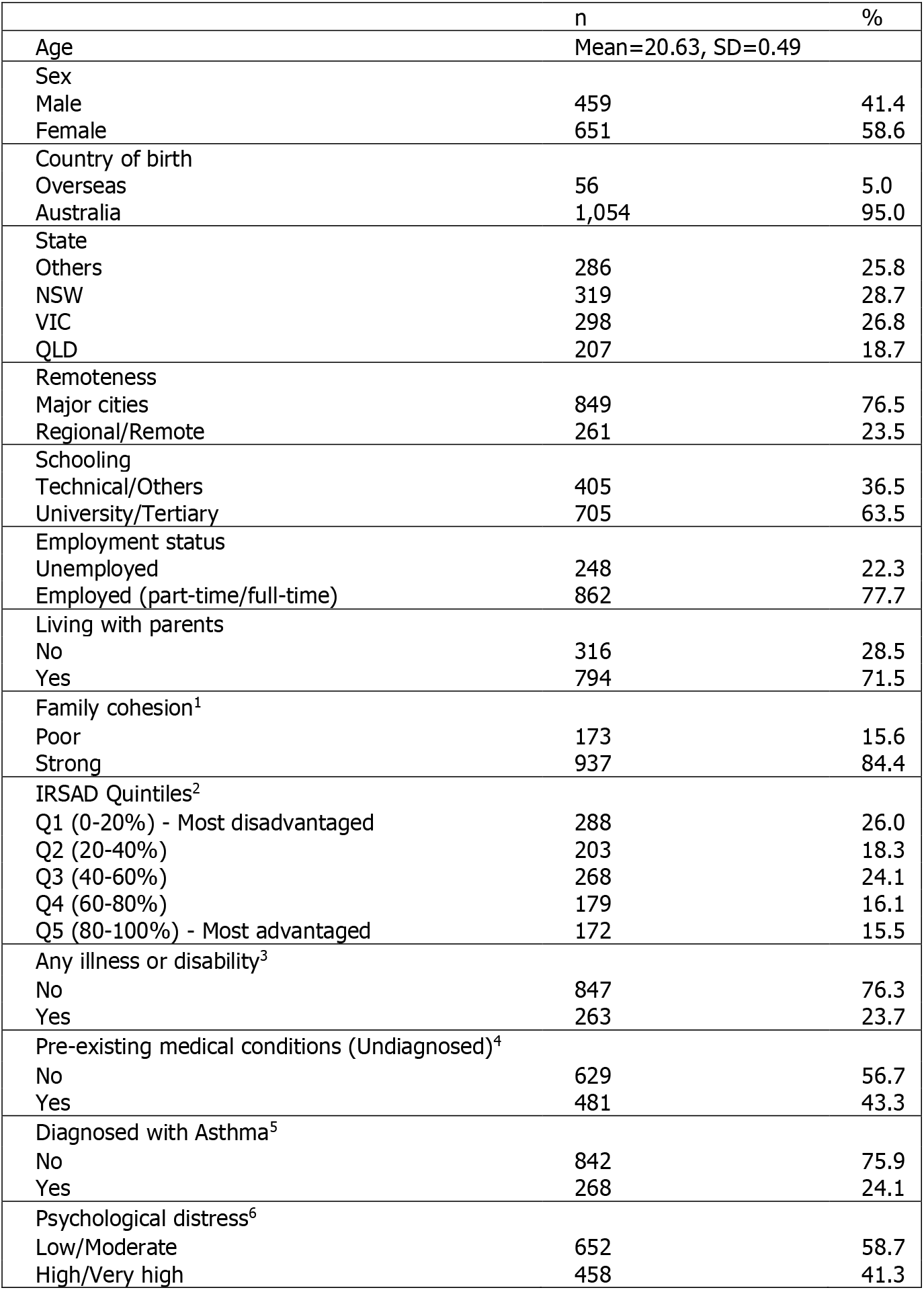
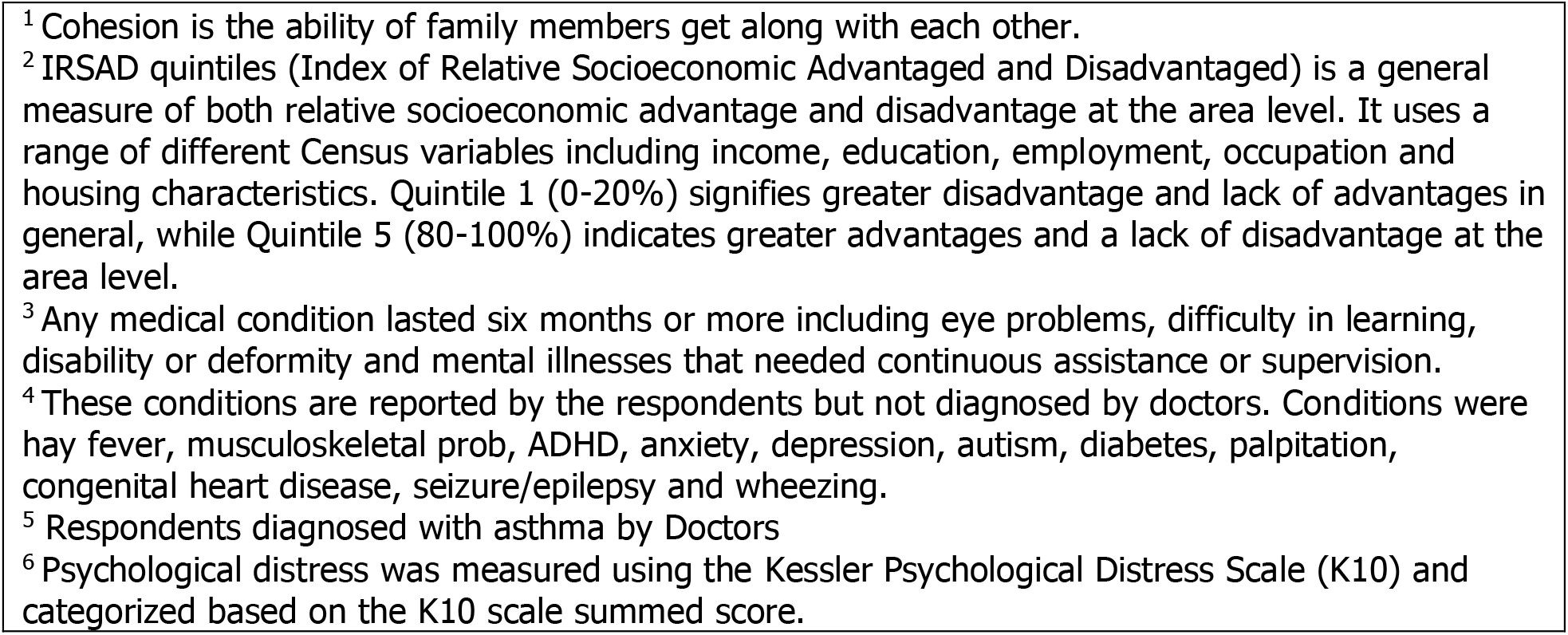
Sample characteristics (n=1110)

|  | n | % |
| --- | --- | --- |
| Age | Mean=20.63, SD=0.49 |  |
| Sex |  |  |
| Male | 459 | 41.4 |
| Female | 651 | 58.6 |
| Country of birth |  |  |
| Overseas | 56 | 5.0 |
| Australia | 1,054 | 95.0 |
| State |  |  |
| Others | 286 | 25.8 |
| NSW | 319 | 28.7 |
| VIC | 298 | 26.8 |
| QLD | 207 | 18.7 |
| Remoteness |  |  |
| Major cities | 849 | 76.5 |
| Regional/Remote | 261 | 23.5 |
| Schooling |  |  |
| Technical/Others | 405 | 36.5 |
| University/Tertiary | 705 | 63.5 |
| Employment status |  |  |
| Unemployed | 248 | 22.3 |
| Employed (part-time/full-time) | 862 | 77.7 |
| Living with parents |  |  |
| No | 316 | 28.5 |
| Yes | 794 | 71.5 |
| Family cohesion<sup>1</sup> |  |  |
| Poor | 173 | 15.6 |
| Strong | 937 | 84.4 |
| IRSAD Quintiles<sup>2</sup> |  |  |
| Q1 (0-20%) - Most disadvantaged | 288 | 26.0 |
| Q2 (20-40%) | 203 | 18.3 |
| Q3 (40-60%) | 268 | 24.1 |
| Q4 (60-80%) | 179 | 16.1 |
| Q5 (80-100%) - Most advantaged | 172 | 15.5 |
| Any illness or disability<sup>3</sup> |  |  |
| No | 847 | 76.3 |
| Yes | 263 | 23.7 |
| Pre-existing medical conditions (Undiagnosed)<sup>4</sup> |  |  |
| No | 629 | 56.7 |
| Yes | 481 | 43.3 |
| Diagnosed with Asthma<sup>5</sup> |  |  |
| No | 842 | 75.9 |
| Yes | 268 | 24.1 |
| Psychological distress<sup>6</sup> |  |  |
| Low/Moderate | 652 | 58.7 |
| High/Very high | 458 | 41.3 |

<sup>1</sup> Cohesion is the ability of family members get along with each other.
<sup>2</sup> IRSAD quintiles (Index of Relative Socioeconomic Advantaged and Disadvantaged) is a general measure of both relative socioeconomic advantage and disadvantage at the area level. It uses a range of different Census variables including income, education, employment, occupation and housing characteristics. Quintile 1 (0-20%) signifies greater disadvantage and lack of advantages in general, while Quintile 5 (80-100%) indicates greater advantages and a lack of disadvantage at the area level.
<sup>3</sup> Any medical condition lasted six months or more including eye problems, difficulty in learning, disability or deformity and mental illnesses that needed continuous assistance or supervision.
<sup>4</sup> These conditions are reported by the respondents but not diagnosed by doctors. Conditions were hay fever, musculoskeletal prob, ADHD, anxiety, depression, autism, diabetes, palpitation, congenital heart disease, seizure/epilepsy and wheezing.
<sup>5</sup> Respondents diagnosed with asthma by Doctors
<sup>6</sup> Psychological distress was measured using the Kessler Psychological Distress Scale (K10) and categorized based on the K10 scale summed score.

Figure 1 depicts the distribution of service access in the whole sample during COVID-19 times and pre-COVID-19 times. About 40% of respondents did not seek medical care in the past 12 months during COVID-19 times in 2020, compared to 42% in pre-COVID-19 times in 2018. Table 2 provides a detailed breakdown of the reasons for those who did not seek medical care during COVID-19, and pre-COVID-19, times. In both times, respondents mostly did not access services either because they thought the problem would resolve, or the problem already resolved. In addition, a significant proportion reported lack of access because they were afraid of doctors or visiting healthcare, this was more pronounced during COVID-19 times.

**Table 2.** Reasons for not seeking medical care

| Reasons |  | Wave 9C1 (n=440)<br>n (%) | Wave 8 (n=460)<br>n (%) |
| --- | --- | --- | --- |
| 1 | Didn't know who to go and see | 71 (16.1) | 47 (10.2) |
| 2 | Had no transportation | 18 (4.1) | 11 (2.4) |
| 3 | No one available to go along with | 16 (3.6) | 11 (2.4) |
| 4 | Difficult to make appointment | 78 (17.7) | 51 (11.1) |
| 5 | Afraid of what doctors would say or do | 116 (26.4) | 84 (18.3) |
| 6 | Thought the problem would go away | 246 (55.9) | 164 (35.7) |
| 7 | Couldn't pay | 65 (14.8) | 47 (10.2) |
| 8 | The problem went away | 120 (27.3) | 82 (17.8) |
| 9 | Too embarrassed | 84 (19.1) | 59 (12.8) |
| 10 | Felt I would be discriminated against | 10 (2.3) | 8 (1.7) |
| 11 | Didn't think they could help me | 78 (17.7) | 53 (11.5) |
| 12 | Services not available in my area | 14 (3.2) | 9 (2.0) |
| 13 | Others | 65 (14.8) | 44 (9.6) |
| During CRP |  |  |  |
| 14 | I did not want to visit doctor during the coronavirus restriction period | 96 (21.8) | - |
| 15 | My doctor did not do non-emergency appointments during the coronavirus restriction period | 15 (3.4) | - |
| 16 | Appointment cancelled or deferred indefinitely because of the coronavirus restriction period | 8 (1.8) | - |
| 17 | Isolating due to the coronavirus restrictions | 12 (2.7) | - |
| 18 | Telehealth appointment was the only option available | 37 (8.4) | - |
| * CRP related data not collected in Wave 8 |  |  |  |
| ** Reasons are not mutually exclusive and respondent had the option not to answer. Here, we only included those who responded 'Yes' to the above-mentioned reasons for not accessing services. |  |  |  |

**Figure 1.**
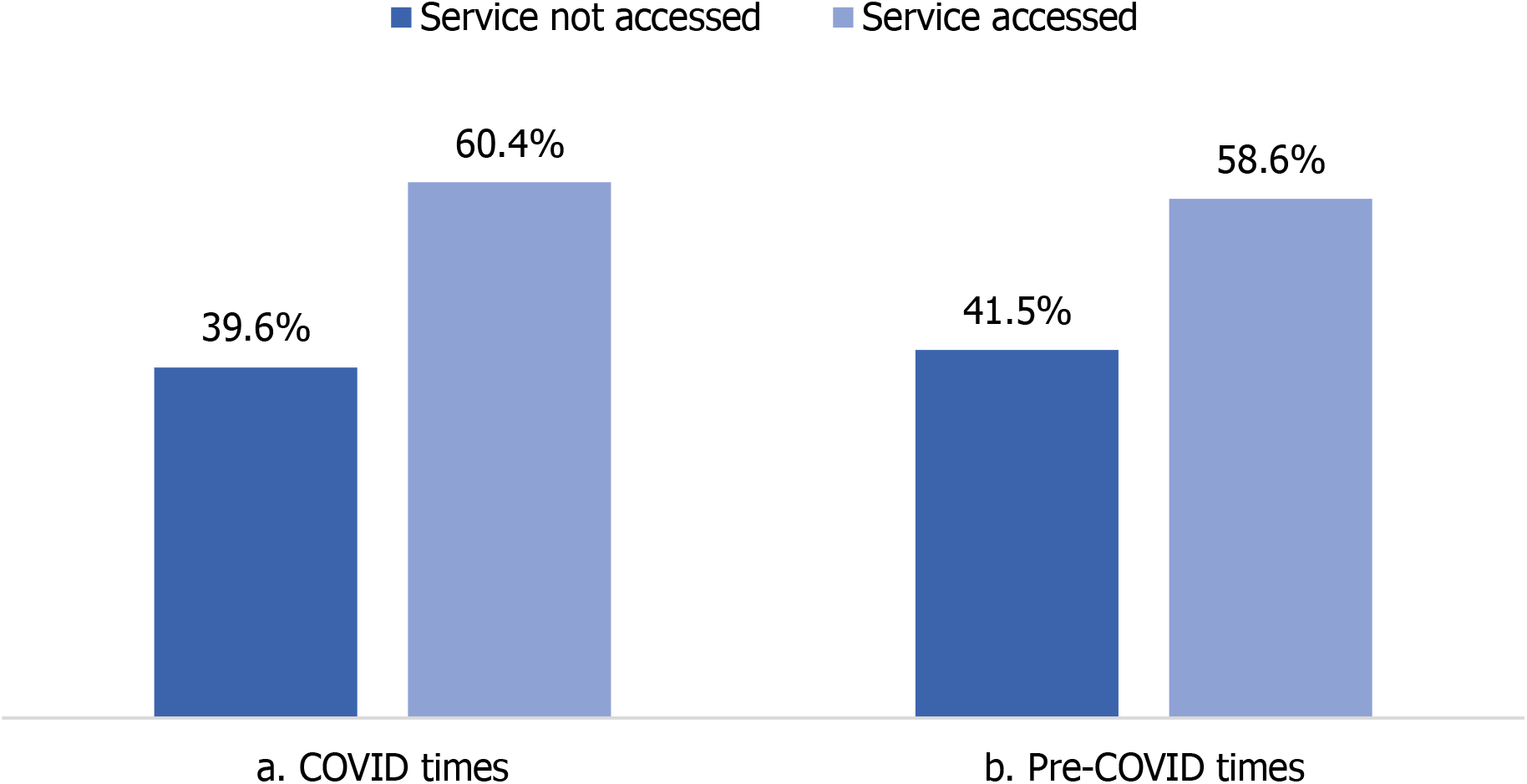
Distribution of access to services - COVID times vs Pre-COVID times a. In between October and December 2020 (during COVID times) respondents were asked whether they seek medical care in the past 12 months for LSAC Wave 9C1. b. In 2018 (pre-COVID times), participants were asked whether they seek any medical care in the previous 12 months for LSAC Wave 8.

The bivariate analysis in Table 3 shows that sex, family cohesion, any illness or disability, self-reported medical conditions, asthma and psychological stress were significantly associated with not seeking healthcare during COVID-19 times. Further, experiencing difficulty of life during COVID-19 times was also significantly associated with not seeking healthcare in the bivariate analysis. While in pre-COVID-19 times, besides these variables, living with one’s parents was also found to be significantly associated with whether an adolescent accessed health services or not

Results from regression models are displayed in Table 4. Model I in Table 4 shows that those who had illnesses or disability that needed continuous assistance and/or supervision were 1.71 times (95% CI: 1.21-2.41) more likely not to access services than those who had no such illnesses or disability during COVID-19 times. Moreover, high/very high psychological stress among youths increased the likelihood (OR 3.17, 95% CI: 2.58-3.91) for not using services compared to those who reported low to moderate psychological stress during COVID-19 times. While in Model II (Table 4), variables associated with barriers to service use during pre-COVID-19 times included respondents who were born in Australia, were from Victoria, had self-reported pre-existing medical conditions, and experienced high to very high psychological stress.

**Table 3.** Factors associated with access to service during COVID times (Wave 9C1) and pre-COVID times (Wave 8) - Bivariate analysis

|  | COVID times<sup>1</sup> (Wave 9C1) |  |  | Pre-COVID times<sup>2</sup> (Wave 8) |  |  |
| --- | --- | --- | --- | --- | --- | --- |
|  | Service not<br>accessed<br>(n=440) | Service<br>accessed<br>(n=670) | Chi-squared<br>test<br>(p-value) | Service not<br>accessed<br>(n=460) | Service<br>accessed<br>(n=650) | Chi-squared<br>test<br>(p-value) |
| Sex |  |  |  |  |  |  |
| Male | 159 (34.6) | 300 (65.4) | 8.18<br>(0.004**) | 173 (37.7) | 286 (62.3) | 4.53<br>(0.033*) |
| Female | 281 (43.2) | 370 (56.8) |  | 287 (44.1) | 364 (55.9) |  |
| Country of birth |  |  |  |  |  |  |
| Overseas | 21 (37.5) | 35 (62.5) | 0.11 (0.737) | 19 (33.9) | 37 (66.1) | 1.37 (0.242) |
| Australia | 419 (39.8) | 635 (60.2) |  | 441 (41.8) | 613 (58.2) |  |
| State |  |  |  |  |  |  |
| Others | 118 (41.3) | 168 (58.7) | 1.15 (0.764) | 110 (38.5) | 176 (61.5) | 2.51 (0.472) |
| NSW | 121 (37.9) | 198 (62.1) |  | 136 (42.4) | 183 (57.4) |  |
| VIC | 115 (38.6) | 183 (61.4) |  | 132 (44.3) | 166 (55.7) |  |
| QLD | 86 (41.5) | 121 (58.5) |  | 82 (39.6) | 125 (60.4) |  |
| Remoteness |  |  |  |  |  |  |
| Major cities | 340 (40.1) | 509 (59.9) | 0.25 (0.617) | 357 (42.1) | 492 (57.9) | 0.55 (0.458) |
| Regional/Remote | 100 (38.3) | 161 (61.7) |  | 103 (39.5) | 158 (60.5) |  |
| Schooling |  |  |  |  |  |  |
| Technical/Others | 161 (39.8) | 244 (60.2) | 0.00 (0.953) | 174 (42.9) | 231 (57.1) | 0.61 (0.435) |
| University/Tertiary | 279 (39.6) | 426 (60.4) |  | 286 (40.6) | 419 (59.4) |  |
| Employment status |  |  |  |  |  |  |
| Unemployed | 107 (43.2) | 141 (56.9) | 1.64 (0.200) | 104 (41.9) | 144 (58.1) | 0.03 (0.858) |
| Employed (part-time/full-time) | 333 (38.6) | 529 (61.4) |  | 356 (41.3) | 506 (58.7) |  |
| Living with parents |  |  |  |  |  |  |
| No | 139 (44.0) | 177 (56.0) | 3.49 (0.062) | 153 (48.4) | 163 (51.6) | 8.85<br>(0.003**) |
| Yes | 301 (37.9) | 493 (62.1) |  | 307 (38.7) | 487 (61.3) |  |
| Family cohesion |  |  |  |  |  |  |
|  | Service not<br>accessed<br>(n=440) | Service<br>accessed<br>(n=670) | Chi-squared<br>test<br>(p-value) | Service not<br>accessed<br>(n=460) | Service<br>accessed<br>(n=650) | Chi-squared<br>test<br>(p-value) |
| Poor | 94 (54.3) | 79 (45.7) | 18.49<br>($<0.001^{***}$) | 87 (50.3) | 86 (49.7) | 6.62<br>(0.010**) |
| Strong | 346 (36.9) | 591 (63.1) |  | 373 (39.8) | 564 (60.2) |  |
| IRSAD Quintiles |  |  |  |  |  |  |
| Q1 (0-20%) - Most disadvantaged | 112 (38.9) | 176 (61.1) | 6.26 (0.180) | 117 (40.6) | 171 (59.4) | 5.38 (0.250) |
| Q2 (20-40%) | 82 (40.4) | 121 (59.6) |  | 81 (39.9) | 122 (60.1) |  |
| Q3 (40-60%) | 119 (44.4) | 149 (55.6) |  | 126 (47.0) | 142 (53.0) |  |
| Q4 (60-80%) | 71 (39.7) | 108 (60.3) |  | 73 (40.8) | 106 (59.2) |  |
| Q5 (80-100%) - Most advantaged | 56 (32.6) | 116 (67.4) |  | 63 (36.6) | 109 (63.4) |  |
| Any illness or disability |  |  |  |  |  |  |
| No | 298 (35.2) | 549 (64.8) | 29.67<br>($<0.001^{***}$) | 333 (39.3) | 514 (60.7) | 6.65<br>(0.010**) |
| Yes | 142 (54.0) | 121 (46.0) |  | 127 (48.3) | 136 (51.7) |  |
| Pre-existing medical conditions<br>(Undiagnosed) |  |  |  |  |  |  |
| No | 229 (36.4) | 400 (63.6) | 6.33<br>(0.012*) | 233 (37.0) | 396 (62.9) | 11.57<br>(0.001**) |
| Yes | 211 (43.9) | 270 (56.1) |  | 227 (47.2) | 254 (52.8) |  |
| Diagnosed with Asthma |  |  |  |  |  |  |
| No | 319 (37.9) | 523 (62.1) | 4.48<br>(0.034*) | 335 (39.8) | 507 (60.2) | 3.93<br>(0.047*) |
| Yes | 121 (45.2) | 147 (54.8) |  | 125 (46.6) | 143 (53.4) |  |
| Psychological distress |  |  |  |  |  |  |
| Low/Moderate | 186 (28.5) | 466 (71.5) | 81.54<br>($<0.001^{***}$) | 220 (33.7) | 432 (66.3) | 38.59<br>($<0.001^{***}$) |
| High/Very high | 254 (55.5) | 204 (44.5) |  | 240 (52.4) | 218 (47.6) |  |
| COVID tested<sup>1</sup> |  |  |  |  |  |  |
| Yes | 129 (40.1) | 193 (59.9) | 0.03 (0.854) | - | - | - |
| No | 311 (39.5) | 477 (60.5) |  |  |  |  |
|  | Service not accessed<br>(n=440) | Service accessed<br>(n=670) | Chi-squared test<br>(p-value) | Service not accessed<br>(n=460) | Service accessed<br>(n=650) | Chi-squared test<br>(p-value) |
| Physical activity during CRP<sup>2</sup> |  |  |  |  |  |  |
| No | 160 (43.7) | 206 (56.3) | 3.79 (0.051) | - | - | - |
| Yes | 280 (37.6) | 464 (62.4) |  |  |  |  |
| Employment status during CRP |  |  |  |  |  |  |
| Unemployed | 285 (38.1) | 464 (61.9) | 2.43 (0.119) | - | - | - |
| Employed (part-time/full-time) | 155 (42.9) | 206 (57.1) |  |  |  |  |
| Received coronavirus supplement<sup>3</sup> during CRP |  |  |  |  |  |  |
| No | 266 (38.3) | 429 (61.7) | 1.45 (0.228) | - | - | - |
| Yes | 174 (41.9) | 241 (58.1) |  |  |  |  |
| Difficulty of life<sup>4</sup> during CRP |  |  |  |  |  |  |
| Few to many | 318 (45.2) | 386 (54.8) | 24.61<br>($<0.001^{***}$) | - | - | - |
| Less or no | 122 (30.1) | 284 (69.9) |  |  |  |  |

<sup>1</sup> Whether the respondent tested for COVID or not. <sup>2</sup> CRP - Coronavirus Restriction Period, between March and May 2020. <sup>3</sup> Coronavirus supplement was the additional financial support for people on welfare during COVID pandemic from Australian Government <sup>4</sup> How difficult was the life during the CRP. Responses included from no problems and/or stresses to many problems and/or stresses. \* CRP (Coronavirus restriction period) related data not collected in Wave 8 \*\* Row percentages are presented

**Table 4.** Determinants of not using health services during COVID times (Wave 9C1) and pre-COVID times (Wave 8)

|  | Wave 9C1 | Wave 8 |
| --- | --- | --- |
|  | Model I<br>OR (95% CI)<sup>1</sup> | Model II<br>OR (95% CI) |
| Sex |  |  |
| Male | Ref | Ref |
| Female | 1.28 (0.93, 1.78) | 1.17 (0.98, 1.39) |
| Country of birth |  |  |
| Overseas | Ref | Ref |
| Australia | 1.59 (0.82, 3.07) | 1.71* (1.10, 2.64) |
| State |  |  |
| Others | Ref | Ref |
| NSW | 1.18 (0.81, 1.71) | 1.55 (0.92, 2.61) |
| VIC | 1.24 (0.81, 1.91) | 1.73** (1.22, 2.45) |
| QLD | 1.16 (0.73, 1.86) | 1.01 (0.71, 1.43) |
| Remoteness |  |  |
| Major cities | Ref | Ref |
| Regional/Remote | 0.94 (0.55, 1.61) | 0.76 (0.42, 1.37) |
| Schooling |  |  |
| Technical/Others | Ref | Ref |
| University/Tertiary | 1.16 (0.90, 1.49) | 0.91 (0.61, 1.35) |
| Employment status |  |  |
| Unemployed | Ref | Ref |
| Employed (part-time/full-time) | 1.04 (0.53, 2.02) | 1.02 (0.75, 1.41) |
| Living with parents |  |  |
| No | Ref | Ref |
| Yes | 0.92 (0.63, 1.34) | 0.65 (0.41, 1.03) |
| Family cohesion |  |  |
| Poor | Ref | Ref |
| Strong | 0.69 (0.46, 1.02) | 0.66 (0.35, 1.22) |
| IRSAD Quintiles |  |  |
| Q1 (0-20%) - Most disadvantaged | Ref | Ref |
| Q2 (20-40%) | 1.38 (0.61, 3.12) | 1.14 (0.91, 1.43) |
| Q3 (40-60%) | 1.24 (0.62, 2.49) | 1.40 (0.89, 2.18) |
| Q4 (60-80%) | 1.18 (0.53, 2.62) | 1.01 (0.72, 1.39) |
| Q5 (80-100%) - Most advantaged | 0.87 (0.32, 2.33) | 0.93 (0.52, 1.67) |
| Any illness or disability (that needs support) |  |  |
| No | Ref | Ref |
| Yes | 1.71** (1.21, 2.41) | 0.97 (0.67, 1.44) |
| Pre-existing medical conditions (Undiagnosed) |  |  |
| No | Ref | Ref |
| Yes | 1.36 (0.98, 1.89) | 1.48* (1.03, 2.12) |
| Diagnosed with Asthma |  |  |
|  | Model I<br>OR (95% CI)<sup>1</sup> | Model II<br>OR (95% CI) |
| No | Ref | Ref |
| Yes | 0.97 (0.61, 1.55) | 1.07 (0.68, 1.69) |
| Psychological distress |  |  |
| Low/Moderate | Ref | Ref |
| High/Very high | 3.17*** (2.58, 3.91) | 2.32*** (1.61, 3.37) |
| COVID tested |  |  |
| Yes | Ref | - |
| No | 1.27 (0.85, 1.89) |  |
| Physical activity during CRP |  |  |
| No | Ref | - |
| Yes | 0.87 (0.65, 1.15) |  |
| Employment status during CRP |  |  |
| Unemployed | Ref | - |
| Employed (part-time/full-time) | 1.13 (0.67, 1.91) |  |
| Received coronavirus supplement during CRP |  |  |
| No | Ref | - |
| Yes | 1.11 (0.68, 1.81) |  |
| Difficulty of life during CRP |  |  |
| Few to many | Ref | - |
| Less or no | 0.84 (0.67, 1.08) |  |
| <sup>1</sup>Adjusted odds ratio (OR) with 95% confidence interval (CI)<br>Level of significance considered: *p<0.05, **p<0.01, ***p<0.001 |  |  |

## DISCUSSION

This study examined factors associated with healthcare access for adolescents and young adults before and during the pandemic. Overall, 39.6% of respondents avoided accessing health services when requiring healthcare, a similar proportion to pre-COVID-19 (41.4%). However, youths with chronic illness or disability, asthma, and those reporting high or very high psychosocial distress perceived significantly greater barriers to healthcare access during the pandemic. Female youths also perceived greater barriers to accessing care.

Young adults with chronic illness or disability felt less able to access services than those without illness or disability. This effect was amplified during COVID-19. This could be attributed to fear of contracting COVID-19 while having a chronic condition or disability, given the greater risk of poorer outcomes in individuals with comorbidities.^28^ Another reason could be the disproportionate side-effects of the COVID-19 pandemic, including changes to travel, isolation requirements and the economic impact including family members losing income or changing employment. Furthermore, adolescents and young adults with chronic illness or disability may have had home environments affected by parental stress and mental ill health during the pandemic. Parents and carers may have experienced increased home demands and decreased support from outside the home, potentially impairing the ability of youths with disability or chronic illness to seek care.^29^

Those diagnosed with asthma, and those with other pre-existing medical conditions, felt less able to access services both before and during COVID-19. This confirms what was known previously, that young people perceive barriers to healthcare access; these barriers were exacerbated by COVID-19.^30^

During COVID-19, females perceived greater barriers to healthcare access than males. Women generally tend to seek healthcare more than men,^31^ suggesting perceived barriers may be higher when desire to access healthcare is higher. However, compared to men, women were more likely to be disadvantaged during the COVID-19 pandemic in that they were more likely to lose their jobs, perform more unpaid labour including carer roles, and be less likely to receive government support,^32^ likely leading to increased stress. Furthermore, an American survey found women attended preventive health services less than men during the pandemic and did not present for recommended medical investigations and treatments.^33^

Youths reporting high family cohesion felt more able to access care when needed compared to those with low family cohesion. This was true both pre- and during COVID-19. Cohesive families may be better at recognising healthcare needs and assisting with the logistical process of attending such healthcare leading to youths being supported and encouraged to attend health services. This support may help to overcome perceived barriers common to young people including cost, transport and waiting times, nervousness and possibly also balancing the need to seek healthcare for a condition while also considering the risk of contracting COVID-19 while attending healthcare facilities.

The majority of young adults with low or moderate psychological distress felt able to access services before (66.3%) and during COVID-19 (71.%). In contrast, the majority of those with high or very high psychological distress felt less able to access services before (52.4%) and during COVID-19 (55.5%). Prior research indicates adolescents and young adults are particularly poor at seeking mental health services,^34^ due to poor mental health knowledge, preference for self-reliance, concerns regarding confidentiality, and lack of resources including money and the availability of professional help.^35^

This study was not without limitations. The sample was not entirely representative of Australian youths; there was an under-representation of Australians born overseas (5% vs. population 30%) and living in rural or remote areas.^36^ Furthermore, most respondents lived at home and reported high family cohesion. Known risk factors for poorer health outcomes include youths living out of home and in insecure housing.^37^ Further research could focus on barriers to healthcare access for youths who live out of home and how the COVID-19 pandemic may have impacted this group of young people.

Another limitation is that this paper describes perceived barriers to healthcare use, not actual healthcare use. Those who perceived more barriers to care may still have accessed healthcare on multiple other occasions during the 12-month study period for each wave of data collection, and possibly more than those who did not identify any barriers to healthcare.

In conclusion, a significant proportion of young adults did not seek care when they felt they needed to seek healthcare during and before the COVID-19 pandemic. The COVID-19 pandemic both modified existing barriers to healthcare access for youth and created new barriers.

## Data Availability

The LSAC datasets are openly available through the National Centre for Longitudinal Data (NCLD) Dataverse, which is publicly accessible upon request, available at https://growingupinaustralia.gov.au/data-and-documentation/accessing-lsac-data. It is not permissible for authors to share the unit record data without approval from the Department of Social Services (DSS) and the Australian Institute of Family Studies (AIFS).

## Author contributions

All authors jointly conceived the study. All authors drafted sections and reviewed the final manuscript. MI and AM coded, analysed and presented the data.

## Data sharing statement

The LSAC datasets are available through the National Centre for Longitudinal Data (NCLD) Dataverse, which is publicly accessible upon request, available at https://growingupinaustralia.gov.au/data-and-documentation/accessing-lsac-data. It is not permissible for authors to share the unit record data without approval from the Department of Social Services (DSS) and the Australian Institute of Family Studies (AIFS).

## Ethics statements

Patient consent for publication: not applicable.

Ethics approval and consent to participate: The LSAC study has been ethically approved by the Human Research Ethics Committee of the AIFS, and written informed consent was obtained for all respondents. Further, the authorship team acquired authorisation from NCLD Dataverse to use LSAC data for research and publications.

